# Simulation of COVID-19 Incubation Period and the Effect of Probability Distribution Function on Model Training Using MIMANSA

**DOI:** 10.1101/2020.06.18.20134460

**Authors:** Abhilasha P. Patel, Arpita A. Welling, Vinay G. Vaidya, Padmaj S. Kulkarni

## Abstract

Coronavirus disease 2019 (COVID-19) has infected people all over the world. While scientists are busy finding a vaccine and medicine, it becomes difficult to control the spread and manage patients. Mathematical models help one get a better feel for the challenges in patient management. With this in mind, our team developed a model called Multilevel Integrated Model with a Novel Systems Approach (MIMANSA) Welling et. al (2020). MIMANSA is a multi-parametric model. One of the challenges in the design of MIMANSA was to simulate the incubation period of coronavirus. The incubation period decides when virus-infected patients would show symptoms. The probability distribution function (PDF), when applied to the number of virus-infected cases, gives a good representation of the process of the incubation period. The probability distribution functions can take various forms. In this paper, we explore a variety of PDFs and their impact on parameter estimation in the MIMANSA model. For our experiments, we used Weibull, Gaussian, uniform, and Gamma distribution. To ensure a fair comparison of Weibull, Gaussian, and Gamma distribution, we matched the peak value of the distribution. Our results show that the Weibull distribution with shape 7.7 and scale 7 for 14 days gives a better training model and predictions.

## 1 Introduction

Several works using trend analysis, growth models, etc, have been done on modelling COVID-19 data. One of the models that simulate real-world observation is a MIMANSA, a model developed by Welling et. al (2020). Multilevel Integrated Model with a Novel Systems Approach, MIMANSA is a novel system approach that models the spread of SARS-CoV-2, a virus that causes COVID-19. One of the factors considered in MIMANSA is the distribution of patients during the incubation period.

One of the challenges in developing a simulator for any virus is to simulate the incubation period. In real life, the incubation period indicates that an infected patient may show symptoms any time from day one to the end of the incubation period. Thus, some infected people may start showing symptoms by the 3rd day while others may show the symptoms by the 8th day. There are different options to represent the incubation period in the mathematical world. One may choose to have a random function that emulates uncertainty of the day of symptoms. Another way could be to use the delay function whose argument uses a random number between 1 to the incubation period. However, an elegant way is of course to use a probability distribution function to represent the incubation period.

In MIMANSA, Welling et.al (2020) used the Weibull distribution to enable the simulation of the incubation period. The question remains, why use the Weibull distribution? How about Gaussian distribution or a uniform distribution? This paper addresses the issue of selecting the best possible function to represent the incubation period.

## 2 Literature Review

Several mathematical models simulate COVID-19 cases. They vary from the exponential, Gompertz, Logistics, to the time series analysis approach using the autoregressive moving average (ARMA) models. Villalobos and Mario (2020) use a generalized logistic equation and the Gompertz equation for fitting COVID-19 data from China. Jia et al. (2020) use Gompertz, Bertalanffy, and Logistic equations for modeling the growth of COVID-19.

Many of the available models cannot handle various issues such as lockdown, quarantine, and exposure changes due to the environmental changes. Additionally, they do not help in considering multiple scenarios that arise in the management of COVID-19.

Time series analysis, using the Auto Regressive Moving Average (ARMA) models, has been another popular approach. This approach has been tried by Deb et al. (2020) on COVID-19. Although it gives initial success in prediction, the success does not last long when the growth rate of cases suddenly changes due to either a sudden outbreak or due to a group of people getting together in large numbers. There is no provision for adding new silent carriers inherent in the ARMA model. If there is a sudden increase in the numbers due to unknown external factors, one has to recalculate the entire model.

Fanelli and Piazza (2020) forecasted cases in China, Italy, and France based on the classical SIRD model. In this approach, the Susceptible (S), Infected (I), Recovered (R), and the Dead (D) model, every person who is going to be infected by the virus, falls in one of the four categories.

Majority of the available models do not have the ability to simulate lockdown, exposure rates, quarantine, or spread in clusters. Additionally, these models are not capable of considering multiple scenarios that arise in the management of COVID-19.

## 3 Simulation of Incubation Period using a Probability Distribution Function

Any person infected by the SARS-CoV-2 virus does not show symptoms immediately. The SARS-CoV-2 virus has an incubation period of 14 days. Thus, even if 10 patients are infected in one day, each one would show symptoms at a different time within the incubation period. This is mathematically represented by applying a Probability Distribution Function (PDF) on the internal count of virus-infected patients (VP). We designed a PDF in such a way that most of the patients show symptoms on the 5th, 6th, and 7th. This makes it possible to simulate the slow release of patients as and when their symptoms show up. In Figure 1, we depict the effect of applying a PDF on virus-infected patients, VP (L, t).

**Figure 1.**
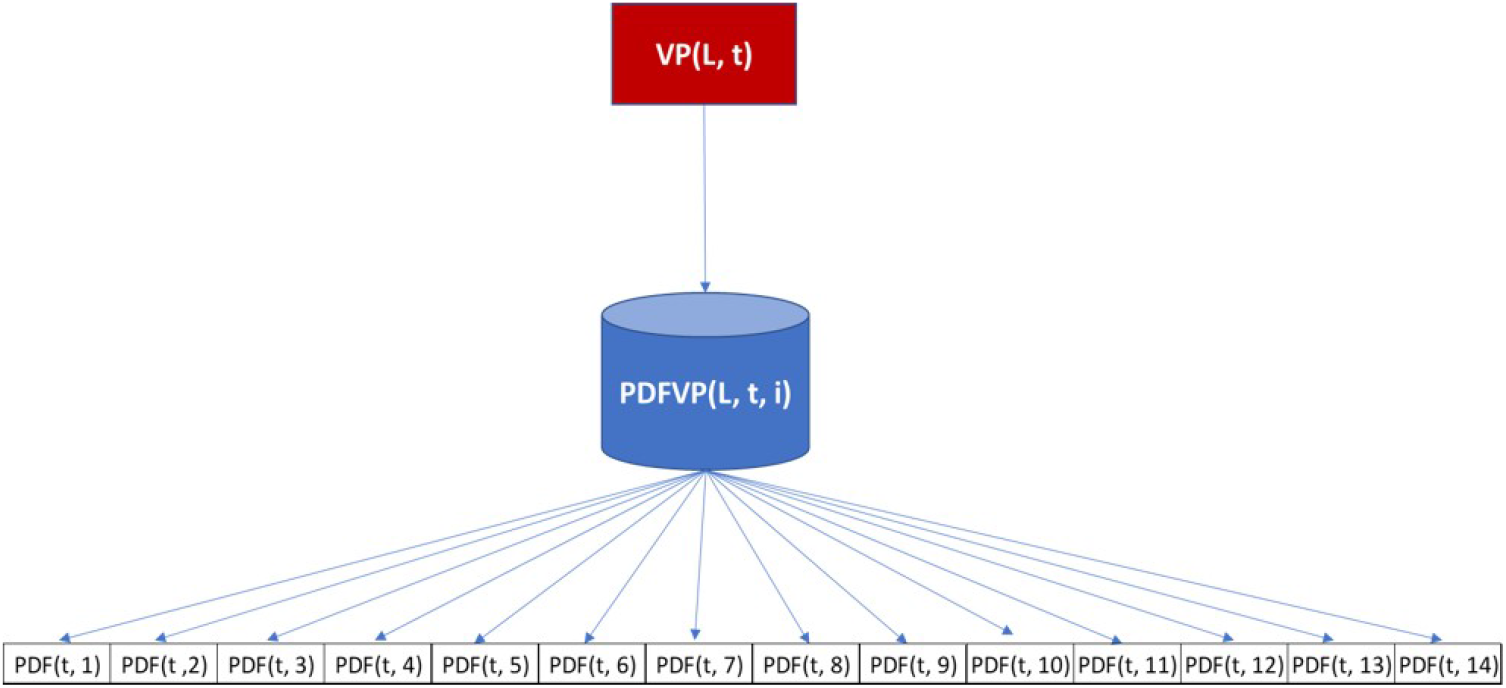
Probability Distribution Function.

The concept of the incubation period is not only applicable to people identified to become virus-infected persons, but it is applicable to silent carriers as well. A person exposed to the virus may exhibit a high rate of infection at any time between 2 to 14 days. It is observed that high activity is around day 5 to 7. Using this observation, we designed another PDF for silent carriers.

## 4 Methods

Welling et. al (2020), has done the parameter estimation so that the optimal trained model is selected using MSE making sure that the parameters that are selected align with the real-world observations. It is observed that the people observe symptoms from day 1 to 14 with symptoms mostly shown on days 5 to 7. To understand the effect of this incubation period we have carried out experiments with most commonly used distributions like Weibull, Gaussian, Gamma, etc. We have also tried to understand how the patient count changes with the change in the pattern when patients are detected early or late. The effect of increase or decrease in the number of patients showing symptoms as per the distribution on patient count is also observed. To see this effect, all parameters are kept the same, only the probability distribution functions are changed. The distributions used for this experiment are:

1. Uniform Distribution
2. Weibull, Gaussian, and Gamma distribution, and
3. Weibull with different Scale and Shape

The following selection criteria were used in finalizing the PDF.

1. Root mean square error (RMSE) between the actual data and the modeled data should be the smallest,
2. Doubling time for the last 10 data points from the actual data should be as close to the doubling time of the modeled data,
3. The slope of the last 10 data points of the actual data should match the slope of the modeled data.

### 4.1 Uniform Distribution

The first uniform distribution that is used is the one that has an equal probability of showing the symptoms throughout the incubation period i.e. 0.07142 for all 14 days. The other distributions have a window of 3 days with value 0.33 that is distributed at other times. For example, 0.33 on day 1,2, and 3, 0.33 on day 2,3, and 4, etc. The Probability density function of the uniform distribution is as in equation 4.1.1 where a and b are the boundaries of the uniform distribution window.The Figure 2 shows uniform distributions considered for simulating the incubation period.

**Figure 2.**
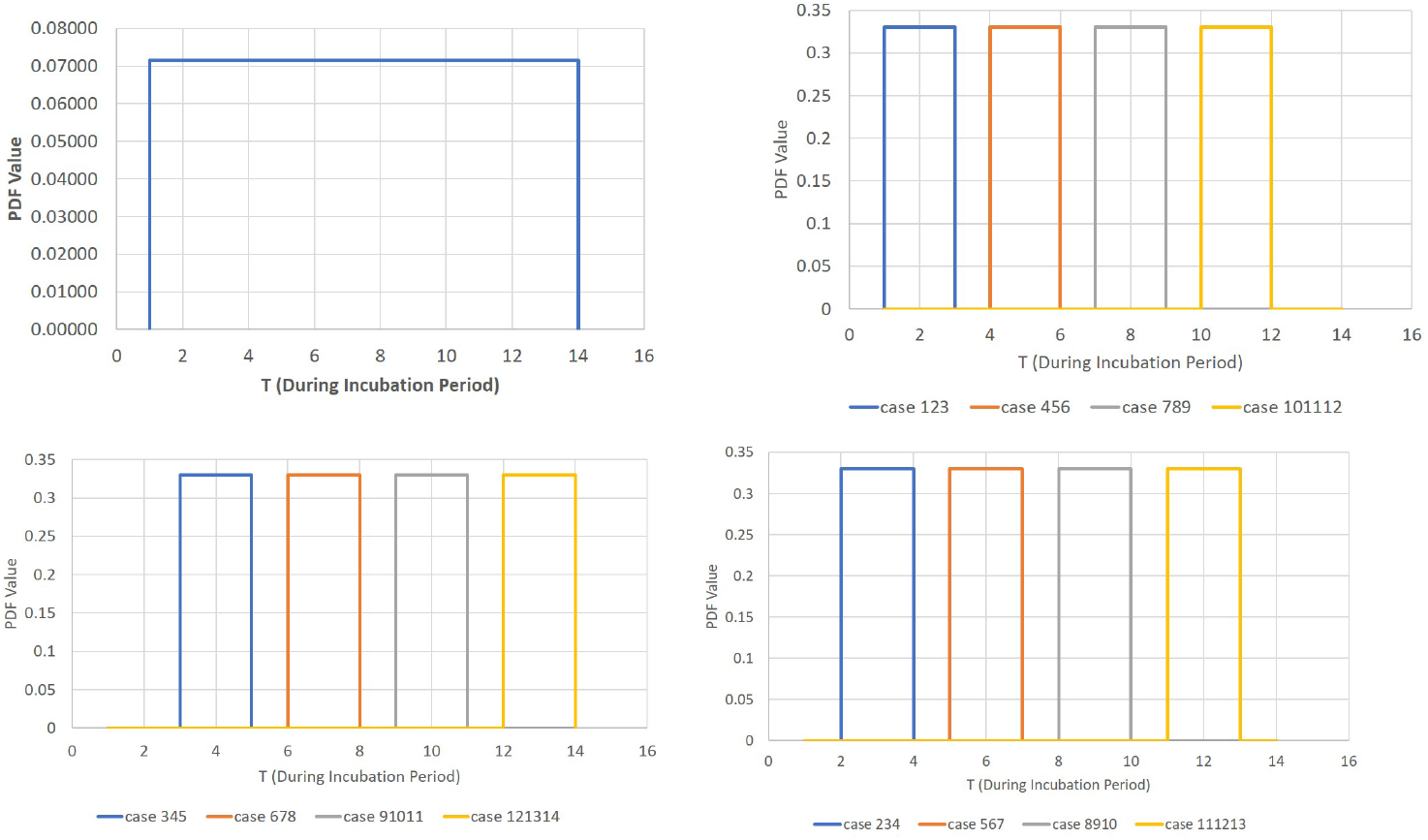
Uniform Distribution with shifting time frame.

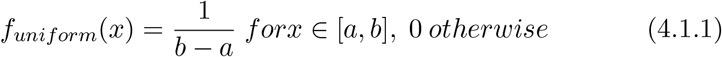

### 4.2 Weibull, Gaussian, and Gamma distribution

Three widely used distributions are Weibull, Gamma, and Gaussian. In this case magnitude of each distribution is kept similar so that any distribution does not have the advantage of having greater magnitude than others. Weibull and Gamma have shapes 3.5 and 6 while scaling 7 and 0.9 respectively. Gamma on the other hand has a mean 8 and standard deviation as 2.1. The probability density functions for each distribution are given by equations 4.2.1, 4.2.2, 4.2.3, and 4.2.4. The graph of the data is shown in Figure 3.

**Figure 3.**
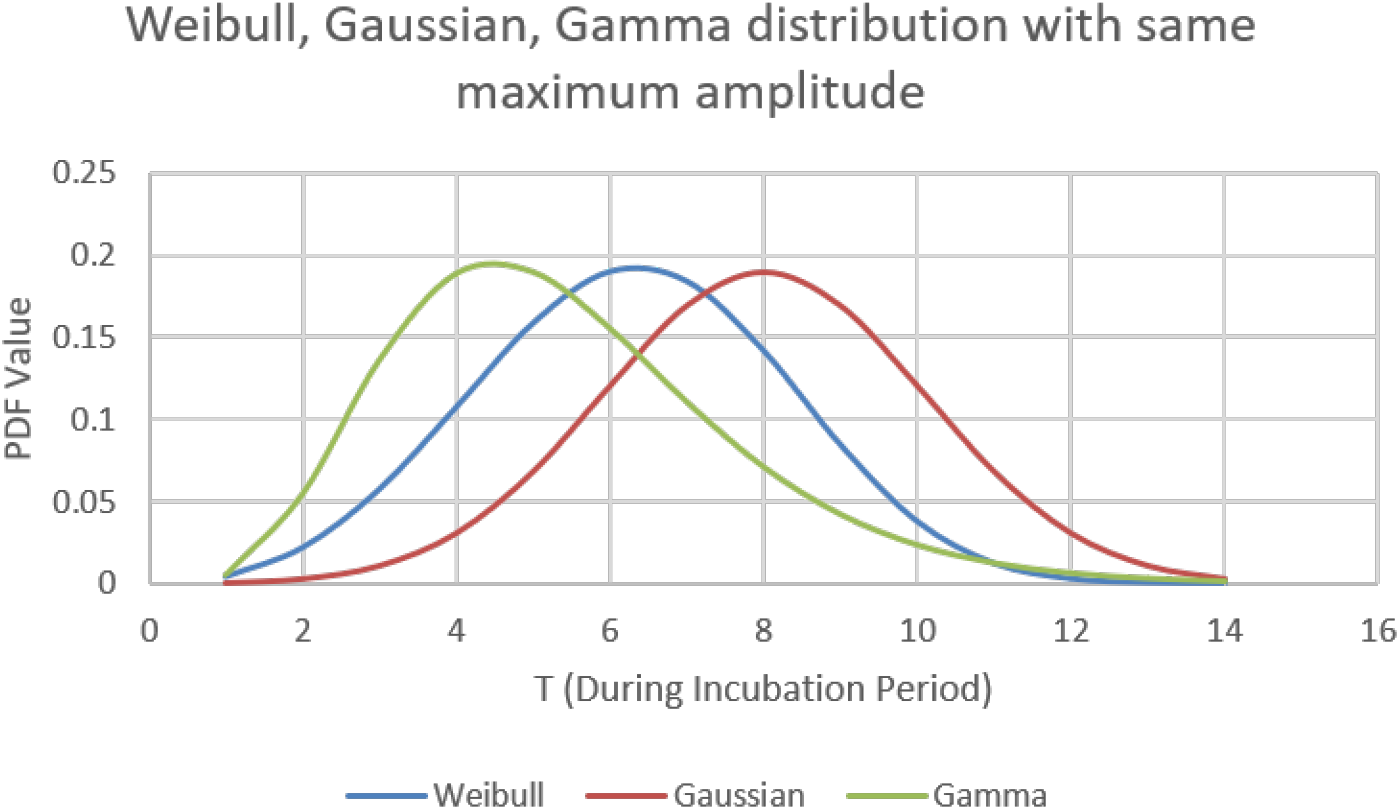
Weibull, Gaussian, and Gamma distribution.

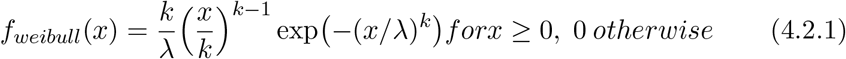

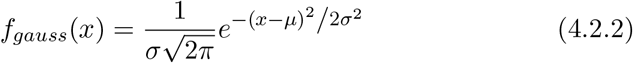

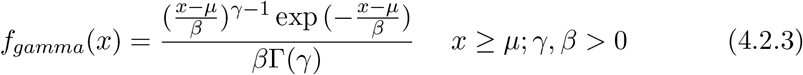

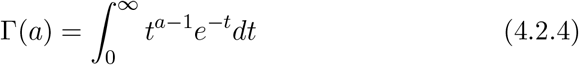

Here, λ = Shape parameter of Weibull distribution, k = Scale parameter of Weibull distribution, σ = standard deviation of Gaussian distribution, µ = Mean of Gaussian and Gamma distribution, β = Scale parameter of Gamma distribution, γ = Shape parameter of Gamma distribution.

### 4.3 Weibull with different Scale and Shape

Due to the versatile nature of Weibull, we dive more into understanding the different nature of Weibull distribution and its effect on the number of people getting affected by COVID-19. Considering the pdf used by Welling et. al (2020) i.e. Weibull (6.3, 8) as a reference of our study, we used Weibull with scale 7,8 and 9 with shape from 2 to 9 for each scale. The best among this was observed and later more analysis with Weibull distribution for more shapes and for the best fit value of scale is carried out. The graph of the data with different shape and scale is shown in Figure 4.

**Figure 4.**
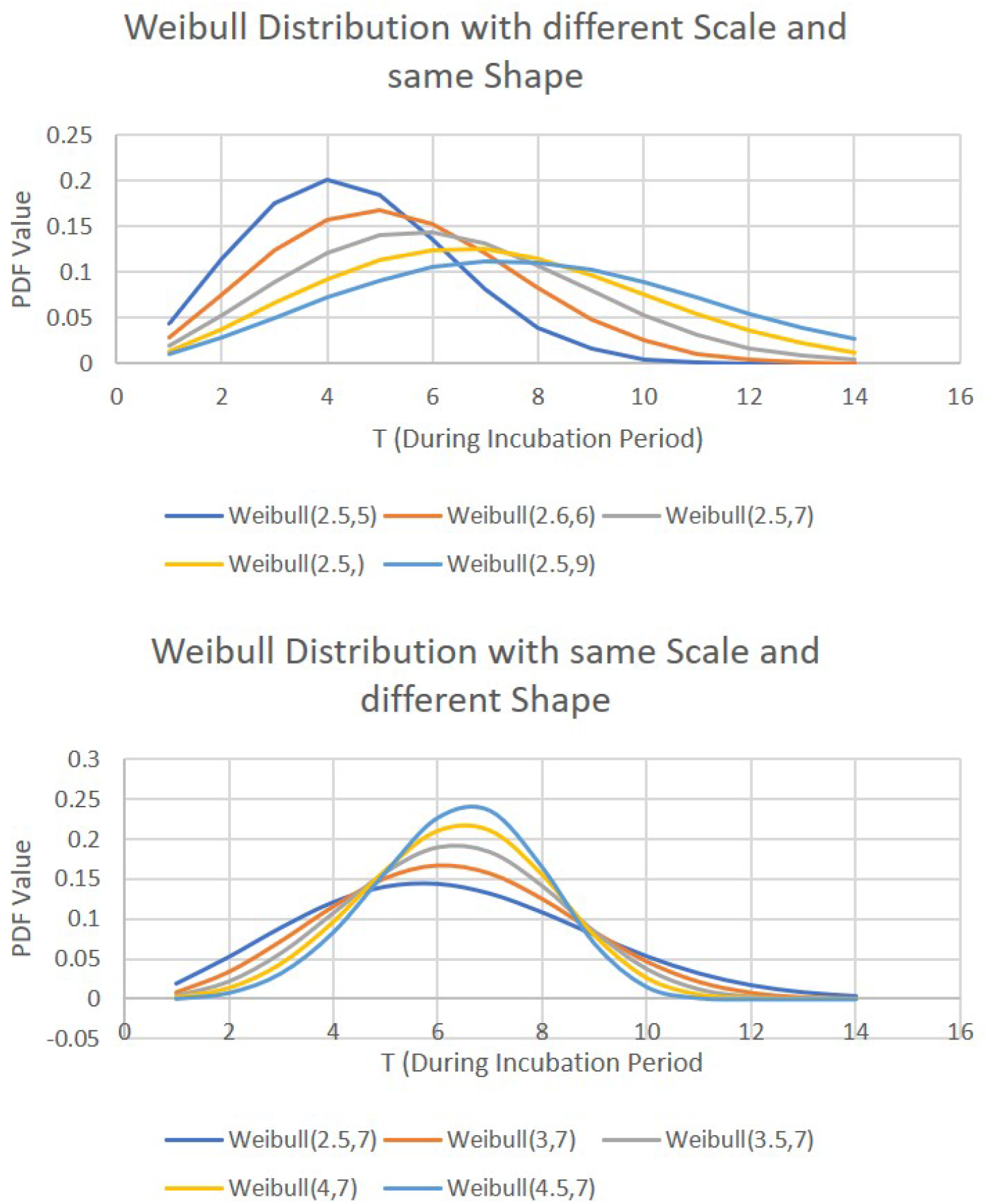
Weibull distribution with different Scale and Shape.

## 5 Results and Discussion

### 5.1 Training with various Distributions

The model was trained using India data from (India data webiste). The model was trained using MIMANSA model with distribution like Weibull, Gaussian, Gamma and Uniform, as discussed in Section 4. Th best case when trained with these distribution is shown in Figure 5 and Figure 6. It is observed as in Figure 5.a that in case of uniform distribution, distribution with equal probablity fits best for India data. Figure 5.b shows how different distributions like Weibull, Gaussian, and Gamma distribution with equal maximum amplitude behaves. It can been seen that Weibull seems to be the best fit. Figure 6.a shows Weibull (8,7) fits best when trained with Weibull having scale 7,8, and 9 with various shapes as discussed in Section 4.3. As in Figure 6.b, it is observed that when data is trained with scale 7 and shapes varying from 2 to 10 with incremental stepsize of 0.1, Weibull(7.7,7) represents the best fit. More details on how these models are selected are discussed in Section 5.2, 5.3, and 5.4.

**Figure 5.**
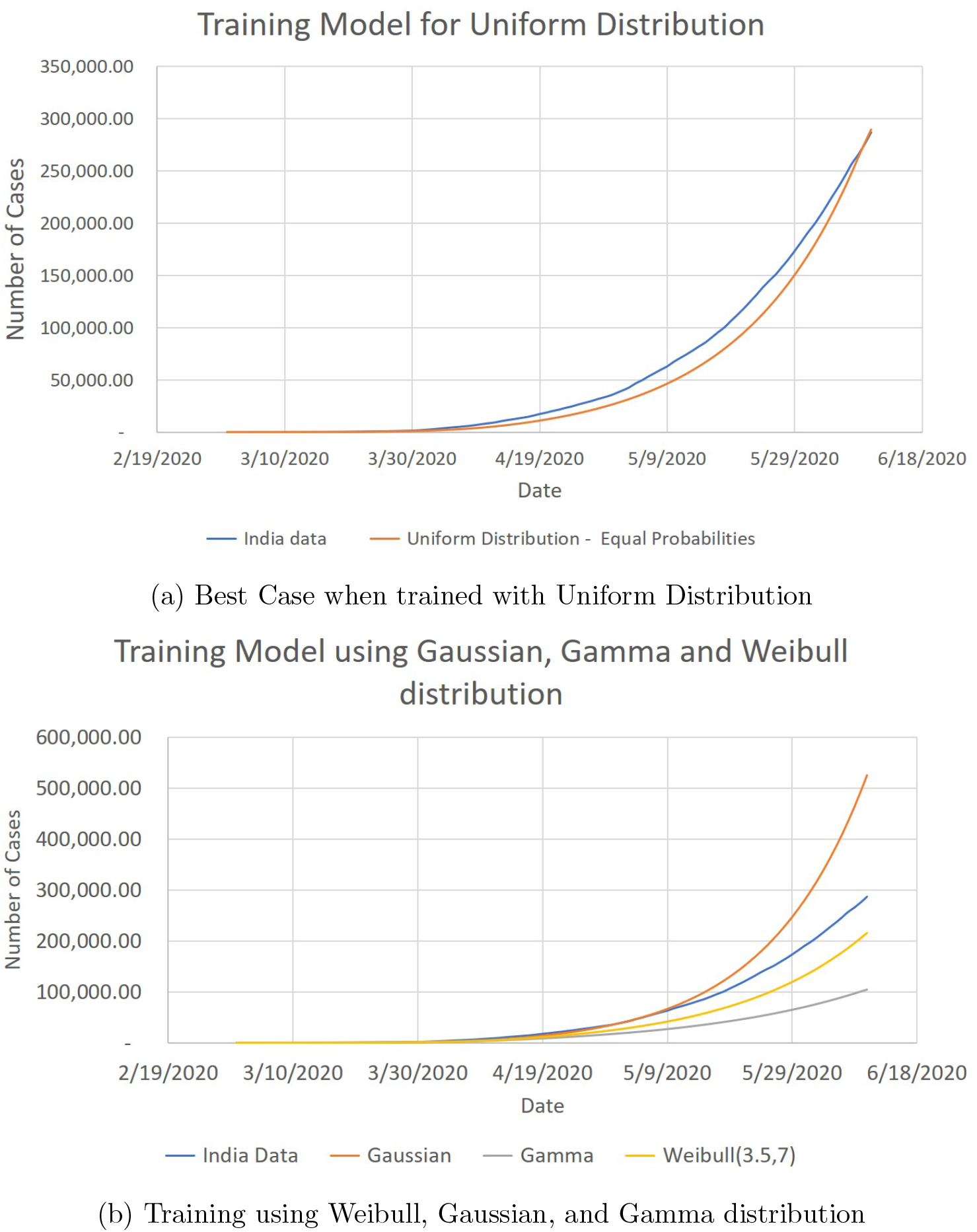
Results of Training using Uniform, Weibull, Gaussian, Gamma Distribution.

**Figure 6.**
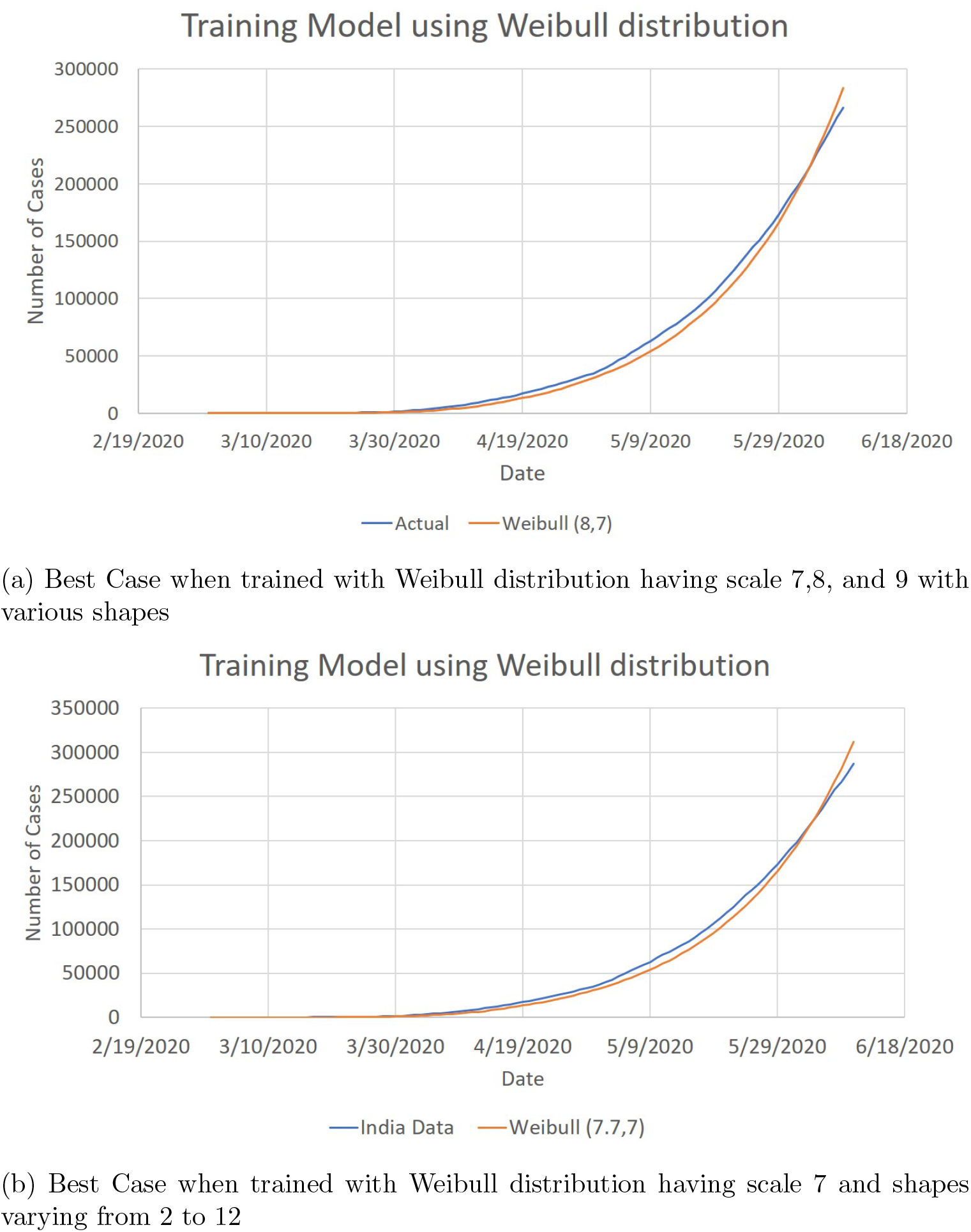
Results of Training using Weibull distribution of different Shape and Scale value.

### 5.2 RMSE as a Criterion for Parameter Estimation

It is common to use the root mean squared error (RMSE) as a criterion for selecting the best model. Figure 7 shows the comparison of the RMSE values for various distributions. In Figure 5, Weibull(S,8) indicates that the Weibull distribution is centered at 8 and takes various shape values, S, ranging from 1 to 12. Similarly, the Weibull(S,7) indicates the Weibull distribution centered at 7. The Weibull RMSE values are plotted in red. The uniform distribution has cases starting with 1 and ending with 12. Each case takes three consecutive values. For example, case 1 indicates the uniform distribution starting at time 1 and has a uniform value for t equal to 1, 2, and 3. The horizontal lines shown in the figure are for the RMSE values for Gaussian, Gamma, and equal distribution.

**Figure 7.**
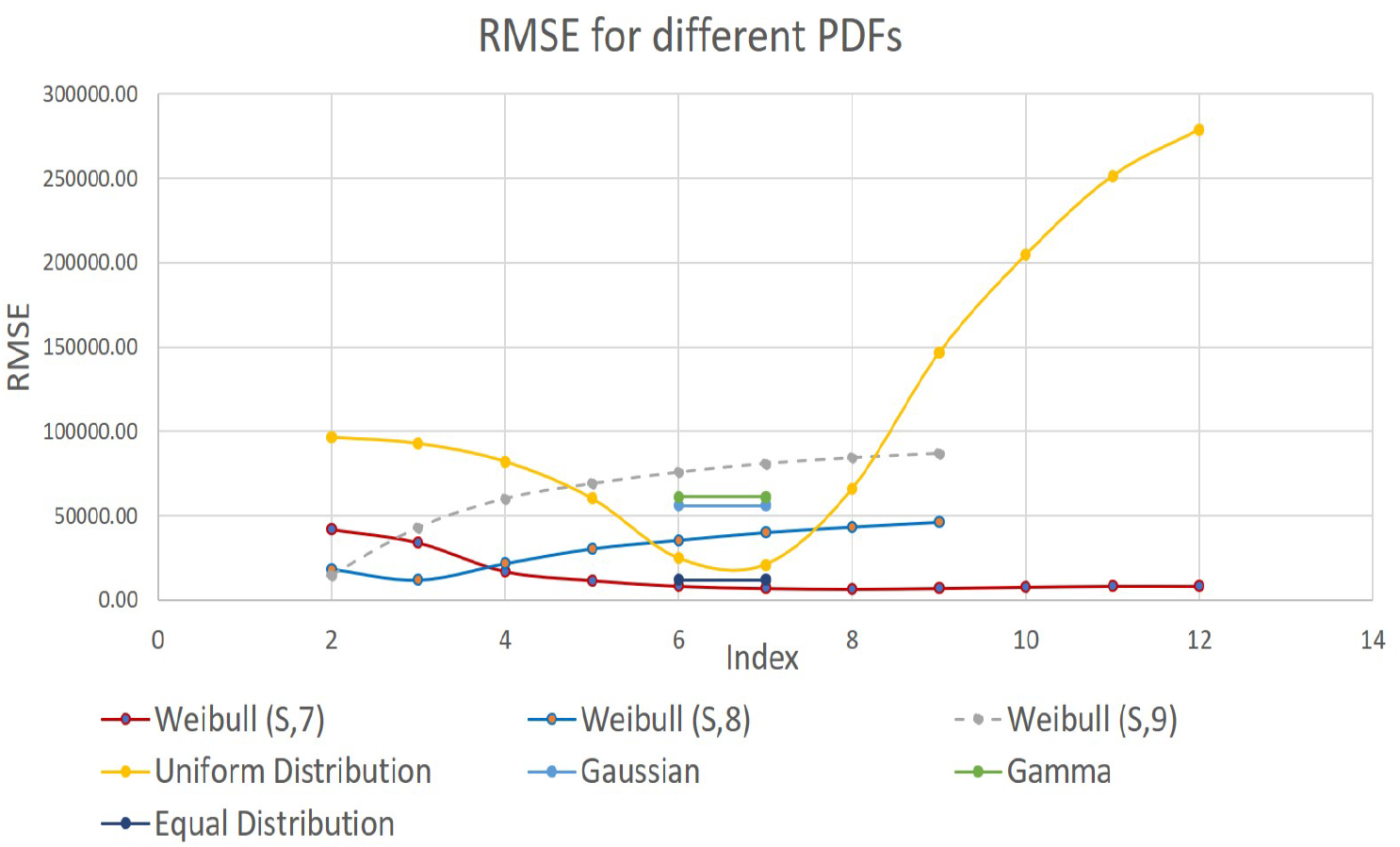
Comparison of RMSE for various distributions.

It is seen that the minimum RMSE value is reached between Weibull(7,7) and Weibull(8,7). At this stage, we do not know the decimal value of the shape factor. Thus, we conducted a study with shape values incremented by starting from 7 and ending at 7.9. Figure 8 shows the variation in the RMSE values when fine-tuning of the shape factor was conducted.

**Figure 8.**
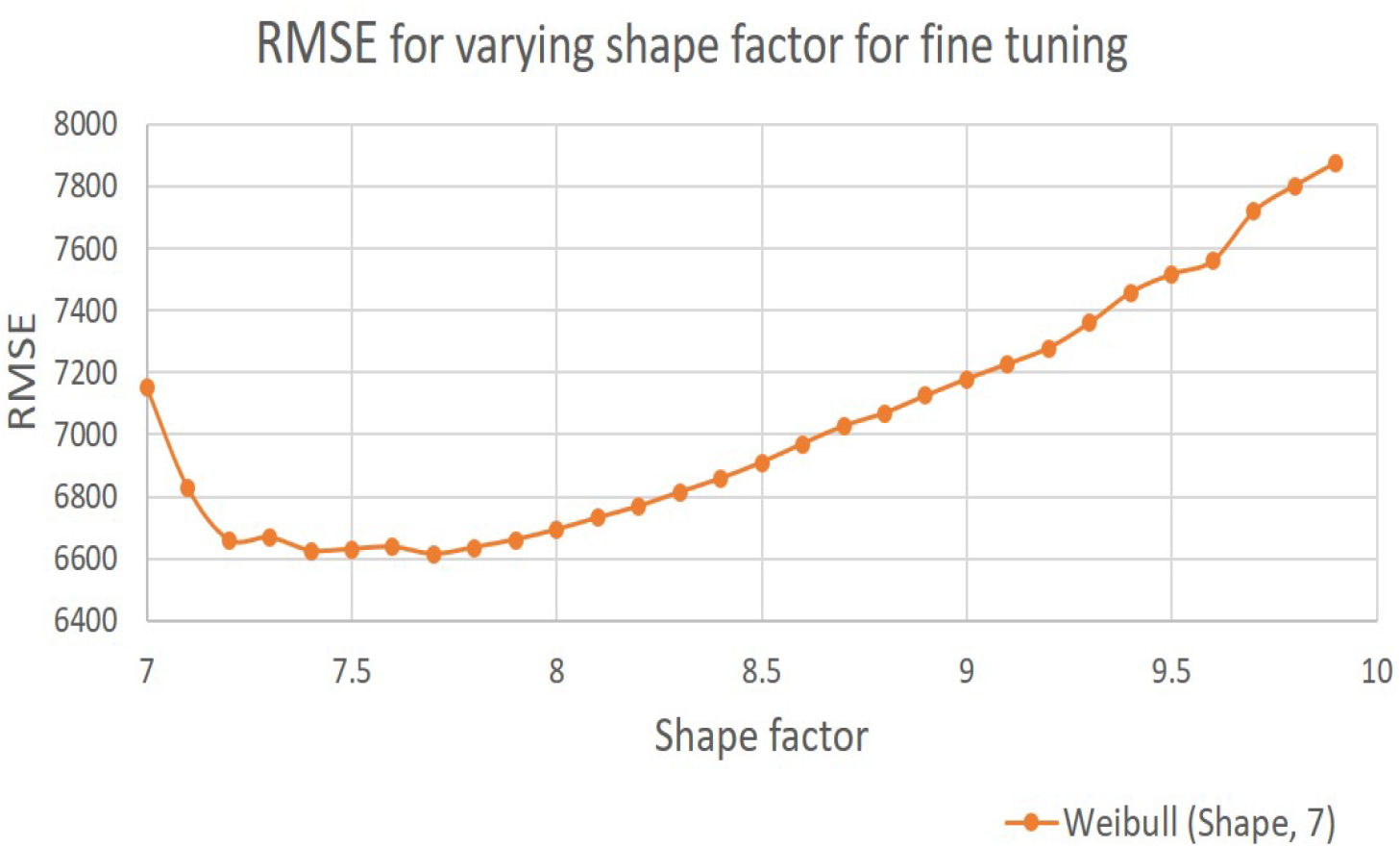
Fine-tuning of the shape parameter value by minimum RMSE.

Figure 8 shows that the minimum value of RMSE is obtained with Weibull distribution with a shape value of 7.7 and a scale value of 7.

It is interesting to note that one would expect the center value of distribution to give the best fit. However, for a uniform distribution, the minimum value is at 6 and for the Weibull distribution, it is at 7.7 for the dataset under investigation. None of the distributions give minima exactly at 7, the midpoint of the 14-day incubation period.

Although we aligned the peak values of Gaussian and Gamma, it did not give the best fit to the COVID-19 data for India.

### 5.3 The Effect of Doubling Time on Selecting the PDF

While studying growth, it is is important to pay attention to the doubling time of growth. The doubling time is defined as the time required for observed data values to double in number. In the case of COVID-19, the number of days it takes for the COVID cases to double is the doubling time. The more the doubling time the better it is since the additional time allows hospitals and administrators to prepare for more cases.

While training the MIMANSA model, finding the least RMSE is only the first step. The training of the model is inadequate if one has the least RMSE with a high error in the doubling time. Such a model will not be able to predict accurate numbers in the future. Thus, we considered the 10 most recent data points to compute the doubling time from India data. This doubling time was compared with the doubling time from various models created by using different PDFs. Figure 9 shows the error in doubling time for various PDFs.

**Figure 9.**
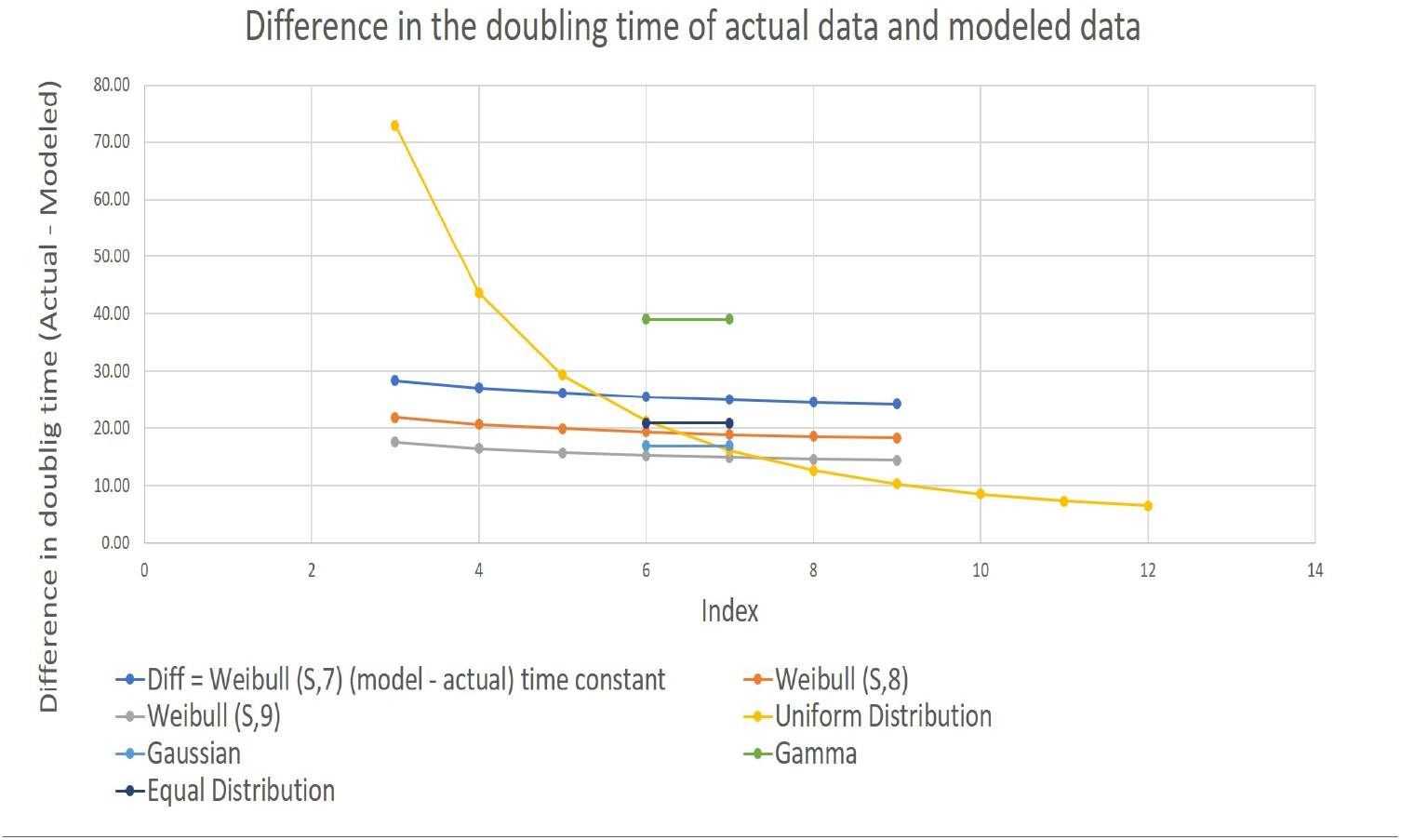
Error in doubling time as a PDF selection criterion.

### 5.4 The Importance of Slope on Future Trend Predictions

Despite using the two criteria such as the least RMSE, and the least error in doubling time, one may still go wrong in predictions. This could be due to the slope of the last 10 data points being different than the slope of the corresponding data points from the actual data set.

Although the slope is related to the doubling time, it gives a different focus on the data trend, and thus it is worth focusing on it.

In Figure 10, we see that the error is negative for all PDFs. It means that all of them will underpredict. All one can do is to choose the PDF that gives the least error in slope.

**Figure 10.**
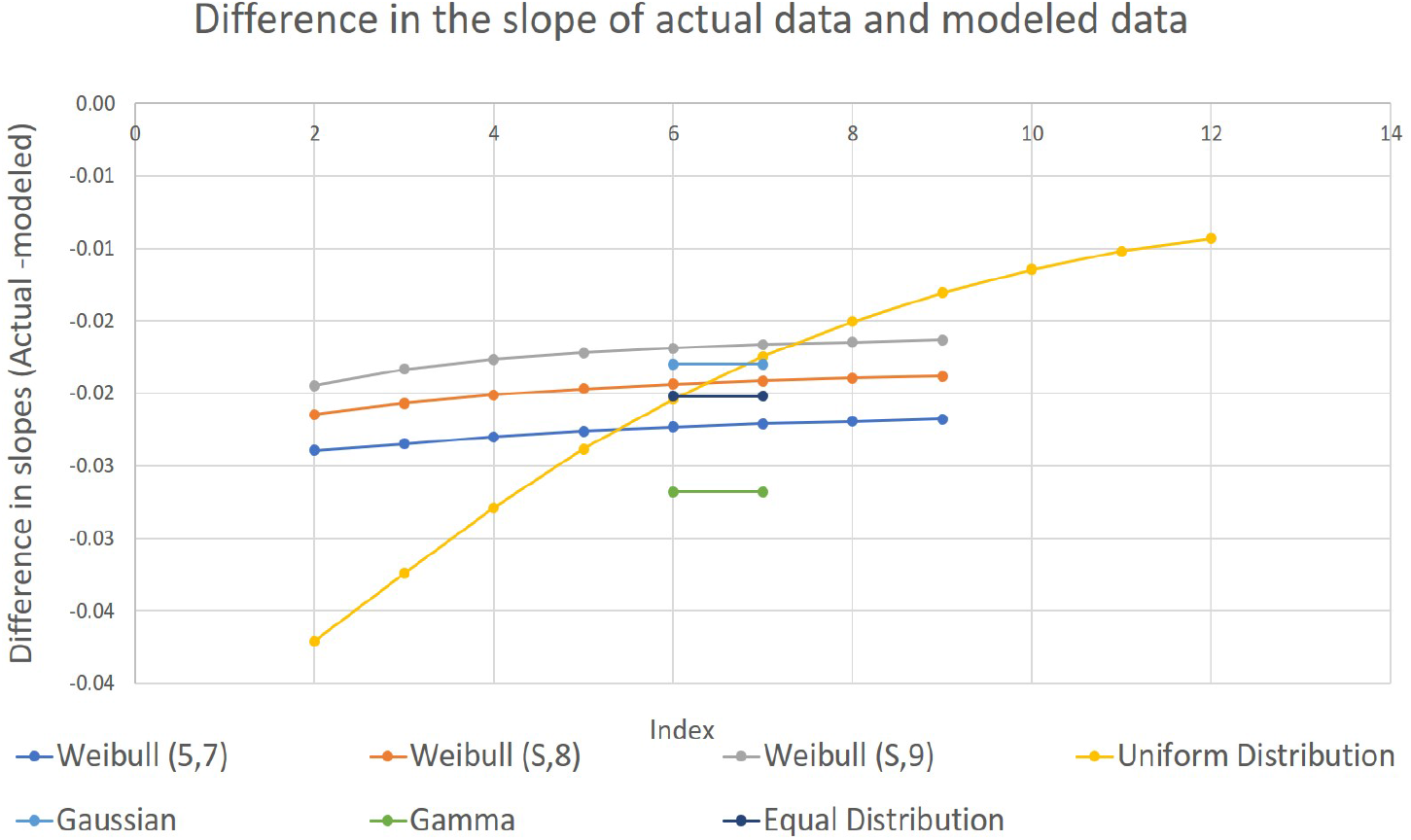
Error in slope as a selection criterionSS.

### 5.5 Selecting the best PDF

As in most situations, selecting the best PDF involves trade-offs. The least RMSE is important to get an overall well-trained model. However, it may compromise the ability to make good predictions in the future. The doubling time is often a yardstick by which growth models are measured. Any compromise in the doubling time may have a serious impact. The slope in the recent past is directly linked with the doubling time. However, when the focus is on the slope, one understands how small numbers may lead to over or under predictions.

In our present case, we found that the Weibull distribution with shape factor 7.7 and scale factor 7 gave a good fit. However, it does lead to error in doubling time and a negative error in slope indicative of under-prediction in the future.

## 6 Conclusion

In the current scenario of the COVID-19 pandemic, there is a need for building a mathematical model of COVID-19 spread. Such a model should have a way to simulate the incubation period of SARS-CoV2. In this paper, we have discussed how to simulate the incubation period using a probability distribution function. Having decided the general methodology, it poses new challenges such as the type of distribution and the parameters of the PDF that would give the best results not only during the training phase of the model but more so during the usage of the model to predict the COVID-19 spread.

The study presented in this paper involved running over 70 different combinations of PDFs and analyzing the generated data. This study used data related to the number of COVID-19 cases in India.

The study concluded that while training the model to a given dataset, three factors determine the best type of PDF to simulate the incubation period of the SARS-CoV2. The factors are the root mean square error, error in doubling time, and the error in the slope of the current values in the actual data. One has to choose the best PDF that would balance the three criteria.

For the Indian dataset, we found that Weibull distribution with shape factor equal to 7.7 and scale factor equal to 7, Weibull (7.7, 7), gave the least RMSE. It was also noted that although it was possible to minimize error in the doubling time by selecting the uniform distribution around T = 12, it was far from the best fit for the given data. For Weibull (7.7, 7), the error in slope was negative. This indicates that the model is likely to project numbers below the actual. This reference also gives a good perspective on the projections from the model.

The methodology presented here enables one to simulate one of the complex elements, the incubation period, in improving our understanding of the spread of the coronavirus. It is hoped that this understanding and better predictions would help us control and manage the pandemic.

## Data Availability

covid19india.org

https://www.COVID19india.org/

## References

[1] Deb, S., and Majumdar M. (2020). A time series method to analyze incidence pattern and estimate reproduction number of COVID-19, arXiv preprint 2003.10655.

[2] Fanelli, D., and Piazza, F. (2020). Analysis and forecast of COVID-19 spreading in China, Italy and France, Chaos, Solitons and Fractals, 134, 109761, 29–46.

[3] India data website, https://www.COVID19india.org/

[4] Jia, L., Li, K., Jiang, Y. and Guo, X. (2020). Prediction and analysis of Coronavirus Disease 2019, arXiv preprint 2003.05447.

[5] Villalobos-Arias, M. (2020). Using generalized logistics regression to forecast population infected by Covid-19, arXiv preprint 2004.02406.

[6] Welling, A. A., Patel, A. P., Kulkarni, P. S. and Vaidya, V. G. (2020). Multilevel Integrated Model with a Novel Systems Approach (MIMANSA) for Simulating the Spread of COVID-19, medRxiv.

[7] Wu, Z., and McGoogan, J. M. (2020). Characteristics of and important lessons from the coronavirus disease 2019 (COVID-19) outbreak in China: summary of a report of 72 314 cases from the Chinese Center for Disease Control and Prevention, Jama, 323 (13), 1239–1242.

